# Medication use and physical assaults in the psychiatric emergency room

**DOI:** 10.1101/2021.05.07.21256772

**Authors:** Y. Nina Gao, Matthew Oberhardt, David Vawdrey, Ryan E. Lawrence, Lisa B. Dixon, Sean X. Luo

## Abstract

**Objective:** To evaluate the relationship between medications used to treat acute agitation (antipsychotics, mood stabilizers and benzodiazepines) and subsequent assault incidence in the psychiatric emergency room.

**Methods:** Medication orders and assault incident reports were obtained from electronic health records for 17,052 visits to an urban psychiatric emergency room from 2014-2019. Assault risk was modeled longitudinally using Poisson mixed-effect regression.

**Results:** Assaults were reported during 0.5% of visits. Intramuscular medications (IMs) were administered in 23.3% of visits overall, and predominately administered within the first 4-hours of a visit. IM administration was correlated with assault (IRR=24.2 [5.33, 110.0]), often because IM medication was administered immediately subsequent to reported assaults. Interacted with time, IMs were not significantly associated with reduction in future assaults (IRR=0.700 [0.467, 1.04]). Neither benzodiazepines nor mood stabilizers were associated with subsequent changes to the risk of reported assault. By contrast, antipsychotic medications were associated with decreased assault risk across time (IRR=0.583 [0.360, 0.942]).

**Conclusions:** IM order rates are high relative to overall assault incident risk. Of the three major categories of medications administered commonly in the psychiatric emergency setting, only antipsychotic medications were associated with measurable decreases in subsequent assault risk. Careful weighing of the risks and benefits of medications is encouraged; antipsychotic medication can have a significant side effect burden, and other medications (IMs, benzodiazepines, mood stabilizers) were not associated with subsequent reduction in assault risk in this analysis.

## Introduction

Physical assaults are a persistent problem in hospital psychiatry, causing physical injuries, psychological distress, and increased costs (1–3). Although by some metrics these events are quite rare (1.98 events per 1000 patient days at one acute psychiatric hospital (4)), other analyses suggest this prevalence is unacceptably high. A recent publication by the Joint Commission (2018) highlighted that 75% of 25,000 workplace assaults reported annually to the Occupational Safety and Health Administration occurred in healthcare and social service settings; workers in healthcare settings are four times more likely to be assaulted than workers in private industry and violence-related injuries are four times more likely to cause healthcare workers to take time off from work. Though, the publication recommends, “Leadership should establish a goal of zero harm to patients and staff…” (5), this charge, to make a rare event even rarer, is fraught with challenges.

Previous investigations have made some inroads by clarifying risk factors for in-hospital physical violence, which include younger age (7,8), male sex (7,9), lower socioeconomic status (10), history of violence (10–12), prior psychiatric hospitalization (11), involuntary hospitalization (7,9,10), substance use (7,9,12–14), personality disorders (8,12), psychosis (7–11,13–15), and institutional staffing and the milieu. Alternatively, although medications (especially antipsychotics, benzodiazepines, and mood stabilizers) are a potentially important tool for modifying violence risk, few studies have analyzed the relationship between medication and violence risk (3,9,13,14,16,17). Studies that did report a relationship offered limited conclusions: non-adherence can be a risk factor for violence (15,18,19), clinicians often respond to aggressive or violent behavior by administering medication (11,20,21), violent patients undergo more medication changes (12), and may receive more medication during their hospital course including higher average daily dose of antipsychotics and benzodiazepines, and multiple antipsychotics (4,7,8,12). However, these studies did not address the central clinical question of whether a particular medication, given at particular moment, may be associated with reduced risk of subsequent violence.

The widespread adoption of electronic health records, which capture the sequence of clinical events in granular detail, has created new opportunities to study the relationship between medication administration and subsequent violence risk. We combined data from the electronic health record with incident reports of violent behaviors (actual or attempted physical assault of another person) to examine whether administering a medication was associated with subsequent reductions in violence risk. Specifically, we asked 1) whether there was evidence that ordering an IM would be associated with reduced risk of subsequent assault in the same visit and 2) whether ordering a particular class of medication (antipsychotics, benzodiazepines or mood stabilizers) would be associated with reduced risk of subsequent assault in the same visit. We chose to focus on patients in an emergency department setting because the prevalence of violence in this setting is relatively high (22) and pharmacological interventions are often first line interventions for highly agitated patients (1).

## Methods

### Setting

Data were collected from a psychiatric emergency room at a large, urban hospital system. The psychiatric emergency room is a locked 24-bed unit staffed with psychiatrists, nurses, nurse practitioners, social workers and recreation therapists, located adjacent to the medical emergency room. Regulations allow patients to be held on an involuntary basis for up to 24 hours, or up to 72 hours in an extended observation room.

### Data

Assault incident reports were collected by the clinical team as part of the organization’s routine monitoring and quality improvement procedure. Hospital staff were required to complete an online incident report for all assaults that occurred, which included a free-text description of what happened, when and where it happened, and who was involved. Data from these incident reports with a location in the psychiatric emergency room were collected from January 2014 through December 2019. The range of precipitants and specific behaviors triggering an assault incident report in this setting have been reported elsewhere (23). Because “assault” language can imply intentionality, uni-directionality, and even moral assessment of the person and situation, we opted to use the more neutral term “incidents” when describing the results.

Data from the electronic health record were obtained for patients evaluated in the psychiatric emergency room from January 2012 through December 2019. These electronic health record data included all medication orders that were written during each patient’s visit as well as demographic variables such as the patient’s age, sex and history of assault and when matched to assault incident report data yielded a census of visits from January 2014 through December 2019.

### Analysis

To organize the data, medication orders were tagged by medication name and route of delivery, merged with incident report data and demographic data, and aggregated to the visit id-hour level. Medication classes were assigned using the table in Appendix A. Several patients appeared to have stays in the data inconsistent with the usual timeframe for a psychiatry emergency room visit. Panels were thus truncated at the 95th percentile of events. The resulting dataset describes each visit from the time of presentation, hour by hour, until the point of discharge.

The psychiatric emergency room treats a heterogeneous population (socially, demographically, ethnically, financially, and diagnostically), therefore, a central analytic challenge was how to compare the risk of a rare event across a wide variety of visit types and presentations. The primary tool we used is mixed-effect Poisson regression. Since existing literature indicated there was wide cross-sectional variation in violence risk across patients, we modeled visits as being drawn from a distribution of risk (captured by visit-level random effects), which we then corrected for when looking within each visit to determine when assaults and medication events occur. This method essentially assigned an individualized baseline level of assault risk to each person in the dataset, then modeled how this risk evolved over the course of the visit.

The hour in which either an assault or a medication event occurred was identified. We defined an indicator marking the hours after a medication event occurred, starting in the hour after the medication was ordered, and modeled this indicator interacted with log time. The resulting estimates can be interpreted as a difference-in-difference comparison (e.g. the difference in assault risk before and after a medication is given for a patient, was compared to the difference in assault risk over the same period of time for another patient who did not receive the medication, acknowledging that risk for both patients would tend to decrease with increasing time in the emergency room).

To better understand how assault risk evolves across a visit, we also identify several windows of time: 4-8 hours before the medication order is written, 1-2 hours before, the hour of the medication order, 1-2 hours after the order, 4-8 hours after the order, 8-16 hours after the order, and greater than 16 hours after the order. These windows are of varying size because, in practice, medication orders are often written in quick succession, particularly around the time of an incident. We ran these variables concurrently in regression with one another so that we could observe how the association of medication interventions and incident risk evolves over time (an “event study”).

The study was approved by the Columbia University Institutional Review Board. Analyses were conducted in STATA 16.1.

## Results

### Study Sample

The matched sample included 1,302,140 hours over 17,052 visits for 9,860 unique patients. The median length of stay for a visit was 10.4 hours. Assault incident reports were relatively rare. From 2014-2019, there were a total of 86 matched assault incident reports. Incidents occurred during 0.5% of visits overall and only 1.7% of visits where the patient had a history of assault (the strongest a priori risk factor available from the literature (9,25)). By contrast, IM medication was ordered at least once during 23.3% of visits, and for patients who had a history of assault in any setting, 29.4% received at least one IM order during subsequent visits. Diagnoses were poorly coded in the data, with only 26.4% of all patients having any recorded ICD-9 or ICD-10 psychiatric diagnosis. Patients with an ICD psychiatric diagnosis had higher rates of IM orders (33.5%) and incidents (1.0%), which was not statistically distinguishable from incident rates among the subsets of visits where a psychotic disorder or substance use disorder were diagnosed.

Figure 1 summarizes the time course of aggregated IM orders and incidents. IM medications were most commonly ordered very early during a patient’s evaluation; 25% of all IM orders occurred within the first 4 hours of a patient’s stay. Incidents peaked somewhat later, approximately 20-30 hours from the time of presentation. Subsequently, the risk of an incident and the risk of IM both declined dramatically with the exception of a small, but observable increase in IM order close to the time of departure from the psychiatric emergency room.

**Figure 1.**
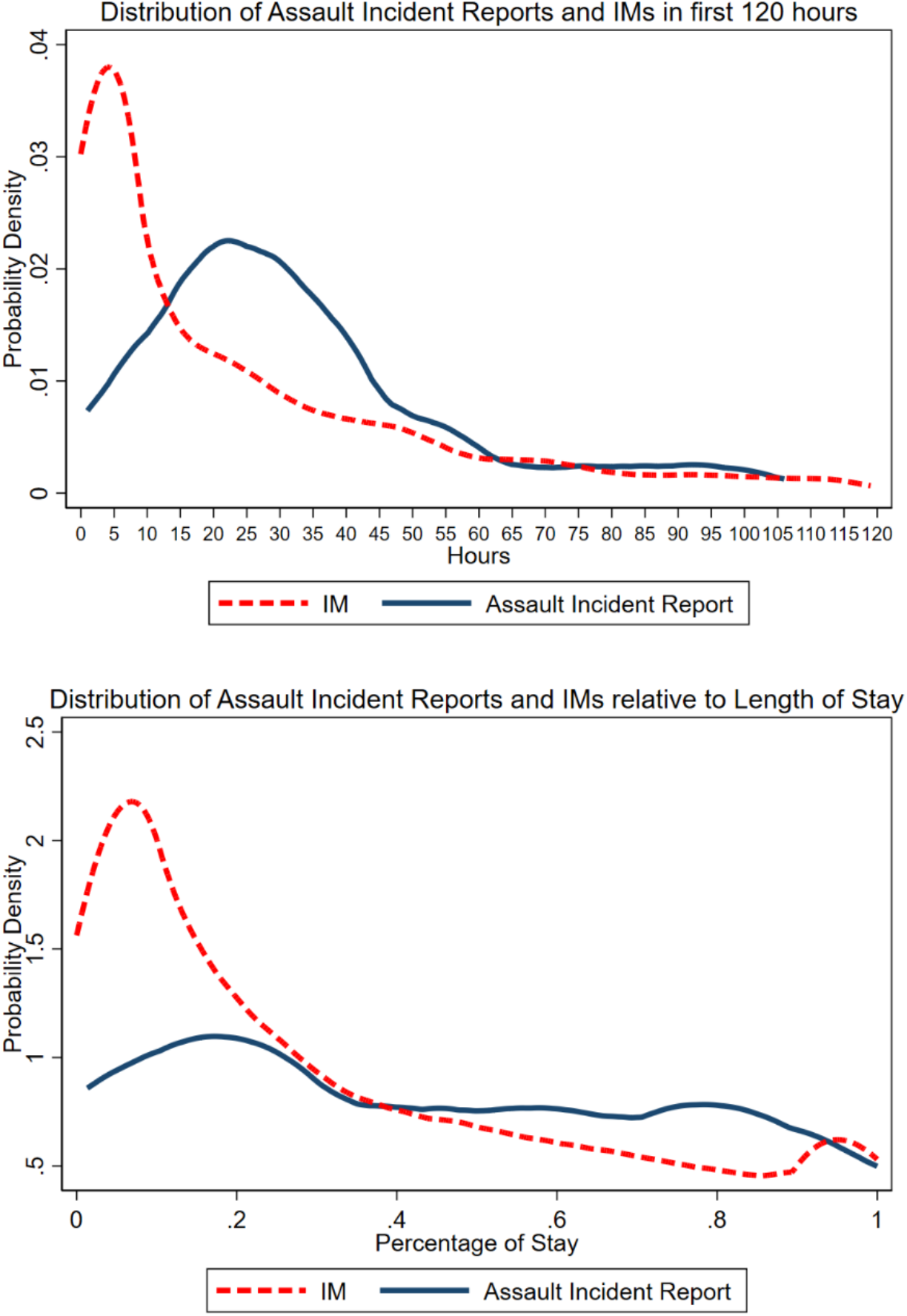
Distribution of Assaults Incident Reports and IMs from the Psychiatric Emergency Room. The distribution of aggregated assault incident reports and IMs across all visits are shown (above) with respect to the first 120 hours in the emergency room and (below) with respect to length of stay for the visits respectively.

Table 2 displays the results from five Poisson mixed-effects regressions. Column 1 pertains to medication route (IM vs. oral). Risk of an incident decreased with time spent in the psychiatry emergency room (IRR=0.425 95% CI [0.311, 0.580]).

**Table 1.** Visit-level variable mean and standard deviation.

|  | **Assault incident reported (N=86)** |  | **No assault incident reported (N=16,970)** |  | **Overall** |  | **T-test** |
| --- | --- | --- | --- | --- | --- | --- | --- |
| Variance | Mean | SD | Mean | SD | Median | Mean | p |
| Female | 0.313 | 0.0503 | 0.380 | 0.00373 | 0 | 0.380 | 0.207 |
| Age | 33. | 1.33 | 40.4 | 0.105 | 39 | 40.4 | <0.001 |
| Length of Stay (Hours) | 251. | 66.5 | 295. | 22.8 | 40.5 | 295. | 0.892 |
| Received IM during visit | 0.813 | 0.0422 | 0.229 | 0.00323 | 0 | 0.233 | <0.001 |
| Received antipsychotic during visit | 0.977 | 0.0163 | 0.673 | 0.00360 | 1 | 0.675 | <0.001 |
| Received benzodiazepine during visit | 0.849 | 0.0388 | 0.751 | 0.00332 | 1 | 0.751 | 0.036 |
| Received mood stabilizer during visit | 0.126 | 0.00255 | 0.267 | 0.0480 | 0 | 0.127 | <0.001 |
| Diagnosis: Any Psychiatric Diagnosis | 0.523 | 0.0542 | 0.262 | 0.00338 | 0 | 0.264 | <0.001 |
| Diagnosis: Primary Psychotic | 0.256 | 0.0473 | 0.108 | 0.00239 | 0 | 0.109 | <0.001 |
| Diagnosis: Depression | 0.0814 | 0.0297 | 0.0852 | 0.00214 | 0 | 0.0851 | 0.901 |
| Diagnosis: Bipolar Disorder | 0.244 | 0.0466 | 0.0856 | 0.00215 | 0 | 0.0865 | <0.001 |
| Diagnosis: Substance Use Disorder | 0.244 | 0.0466 | 0.122 | 0.00251 | 0 | 0.123 | <0.001 |
| Diagnosis: Lack of/Inadequate Housing | 0.0813 | 0.0297 | 0.0464 | 0.00162 | 0 | 0.0467 | 0.126 |
| History of Assault | 0.977 | 0.0163 | 0.284 | 0.00346 | 0 | 0.288 | <0.001 |
| Brought in by EMS | 0.349 | 0.0517 | 0.342 | 0.00364 | 0 | 0.342 | 0.896 |
| Admitted to Inpatient Psych | 0.686 | 0.0503 | 0.651 | 0.00366 | 1 | 0.651 | 0.494 |
| Ever CPEP IM | 0.860 | 0.0376 | 0.356 | 0.00368 | 0 | 0.359 | <0.001 |

Variables summarized at the visit-level. In total, there were 17,056 visits for 9,870 patients with matched data. Means for binary visits represent fraction of visits with that characteristics, for example, 38% of observed visits were for female patients.

**Table 2.** Poisson regressions expressing the relationship of medication administration to incident risk.

|  | (1) | (2) | (3) | (4) | (5) |
| --- | --- | --- | --- | --- | --- |
| Log(Time) | 0.425 **<br>[0.311 0.580] | 0.483 **<br>[0.320 0.732] | 0.426 **<br>[0.314 0.580] | 0.41 **<br>[0.260 0.646] | 0.402 **<br>[0.326 0.495] |
| After IM | 24.2 **<br>[5.33 110.0] |  |  |  |  |
| (After IM)* Log(Time) | 0.700<br>[0.467 1.04] |  |  |  |  |
| After Antipsychotic |  | 14.7 **<br>[3.30 65.4] |  | 26.8 **<br>[4.78 150.4] |  |
| (After Antipsych)* Log(Time) |  | 0.583 **<br>[0.360 0.942] |  | 0.489 **<br>[0.291 0.822] |  |
| After Oral Antipsychotic |  |  | 10.9 **<br>[2.22 53.6] |  |  |
| (After Oral Antipsy)* Log(Time) |  |  | 0.597 **<br>[0.376 0.949] |  |  |
| After Benzodiazepine |  |  |  | 0.342<br>[0.060 1.95] |  |
| (After Benzo)* Log(Time) |  |  |  | 1.49<br>[0.868 2.57] |  |
| After Mood Stabilizer |  |  |  |  | 10.4<br>[0.967 112.8] |
| (After Mood Stab)* Log(Time) |  |  |  |  | 0.635<br>[0.350 1.15] |

The table describes the IRR from four Poisson mixed-effects regressions at the visit-level, each with the outcome of assault. 95% CI given for each variable displayed below. All regressions include age-group controls for ages 30-49, 50-69, and >70 years with age <30 as the reference category. The number of observations was 1,302,140 with 17,052 visits for all of the above. \*\* indicates Wald statistic significant at the 0.05 level.

### Medication Route

The first clinical question we examined was whether ordering an intramuscular medication was associated with reduced risk of subsequent assault in the same visit. Incidents frequently occurred within an hour of IM order, with incidents frequently preceding IM orders (not shown), suggesting that IM medications were more often ordered in response to an incident. Orders for IM medications were associated with an increased risk of subsequently having an incident (Table 2, IRR=24.2 [5.33, 110.0]).

Incident prevention was described by the interaction between that medication and time, also described by Table 2. Here, IM medications were not significantly associated with a reduction in subsequent incidents (IRR=0.700 [0.467, 1.04]).

Figure 2 elaborates on Table 2 results by plotting the estimated Poisson coefficients (i.e., incident risk level) for incidents relative to the timing of medication. Panel A describes the time course for IM medications and shows that, on average, the risk of an incident increases starting one hour before the hour of IM order and peaks during the hour of IM order (point estimate 3.19 [2.56, 3.83]). Subsequently, point estimates for incident risk decrease abruptly, reaching a trough in the 4-7 hours following IM order, which is statistically indistinguishable from baseline risk (−0.116 [-1.19, 0.962]). Among patients who had had an IM ordered earlier in their stay, the risk of having an incident 16-hours post-IM was in fact elevated even relative to the patient’s baseline (point estimates 0.950 [0.190, 1.71]), controlling for time spent in the emergency room.

**Figure 2.**
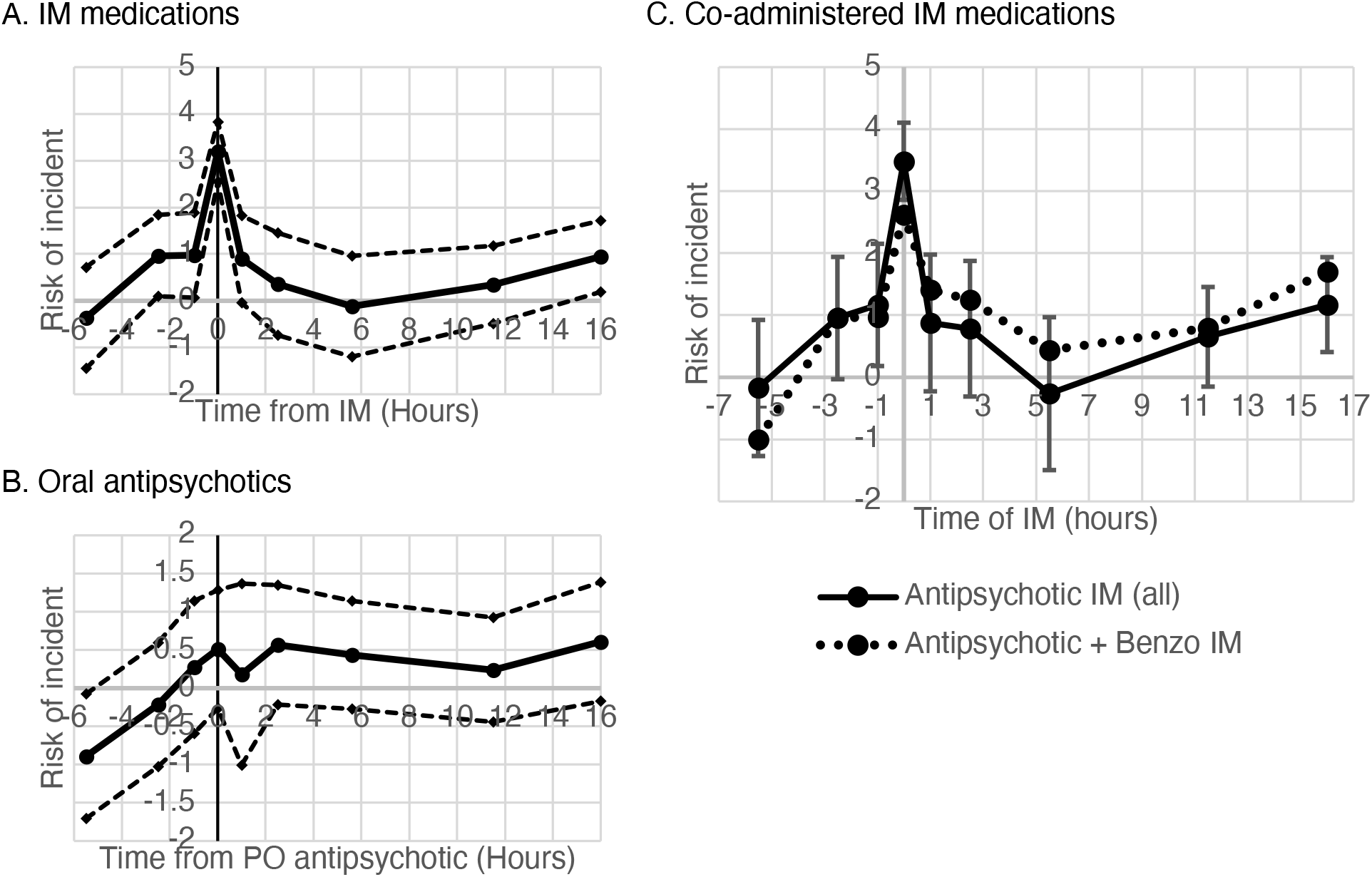
Event study of assault risk relative to timing of medication orders. The figure plots the estimated Poisson coefficients for risk of assault incident report for lagged dependent indicator variables labelled with respect to the hour of medication administration. Coded periods are from 4-8 hours before, from 1-2 hours before the medication order is written, the hour of the medication order, from 1-2 hours after the order, from 4-8 hours after the order, from 8-16 hours after the order and, finally, greater than 16 hours after the order. All regressions include age group and history of IM in CPEP and log(time) controls). Panel A shows event study for all IMs, regardless of medication time and illustrate the extent to which IM medications are administered within the same hour as an assault incident report (elevated risk at time=0), with patients returning to baseline level of risk 4-6 hours after administration. Dashed lines give the 95% confidence interval on estimates. Panels B and C attempt to disaggregate this effect. Panel B displays two curves. The bold curve in gives the aggregated trendline for all IM antipsychotics, which closely resembles panel A. Error bars here give the 95% confidence interval on estimates for IM antipsychotics. The dotted line in panel B gives the trend for only IM antipsychotics coadministered with a benzodiazepine. Panel C displays the results for oral antipsychotics alone, which is modestly downward sloping after time>0. Dashed lines in panel C give the 95% confidence interval on estimates.

### Medication Class

The second clinical question we examined was whether ordering a particular class of medication (antipsychotic, benzodiazepine, or mood stabilizer) was associated with reduced risk of subsequent assault in the same visit. Because antipsychotic medications were the most commonly ordered IM medication in this sample, it was not surprising that antipsychotic medications (Table 2, column 2) were also associated with elevated assault risk after the time of initiation (IRR=14.7 [3.30, 65.4]). However, interacted with time, they were associated with decreased assault risk across time (IRR=0.583 [0.360, 0.942]).

Given that IM medications were not associated with subsequent incident risk reduction, the effect of antipsychotics should derive partially from orally administered medications. Table 2 column 3 confirms this, by isolating oral antipsychotics. Interacted with time, oral antipsychotic orders were associated with incident risk reduction (IRR=0.597 [0.376, 0.949]) comparable for the rate for antipsychotic orders overall. Figure 2, panel B describes a modestly downward slope in incident risk associated with oral antipsychotics up to 16-hours, though incident risk remains statistically indistinguishable from baseline throughout the post-administration period.

Benzodiazepines are frequently co-ordered with antipsychotics for the management of agitation, however, we did not uncover an association between the addition of benzodiazepines and additional incident risk reduction. Figure 2, panel C addresses a hypothesis that, in the setting of an acute escalation in assault risk, the addition of a benzodiazepine to an antipsychotic can accelerate de-escalation within the window of a few hours of administration. However, on investigation (Panel C), no shorter-run risk reductions were observed associated with adjunct benzodiazepine orders.

Similar to benzodiazepines, we did not observe reduced incident risk with mood stabilizers.

## Discussion

In this study, we used a large single site electronic health record database harmonized with assault incident reports to address whether IM medications, antipsychotics, benzodiazepines, or mood stabilizers, ordered in the psychiatry emergency room setting, were associated with a subsequent reduction in the risk of physical assault during that emergency room encounter.

We found that IM medication orders were common (23.3% of visits) while assault incident reports were rare (0.5% of all visits, 1.7% of visits where the patient had a history of assault). Our findings raise the question of whether existing research on risk factors for assault, combined with a regulatory emphasis on violence prevention and “zero harm,” have had an unintended consequence of lowering the threshold for ordering IM medication. While IM medications are an important tool, they come with a complex set of risks, benefits, and implications. Administration of IM medications can increase scene safety in some respects, but--particularly when IM medication is given involuntarily--patients may experience a loss of autonomy, they may experience the intervention as unnecessarily invasive, and the experience may disrupt their relationships with healthcare providers (26,27). Administration of IMs may also temporarily escalate the situation and place the patient and staff members at increased risk of injury.

Furthermore, our data did not show a clear association between IM medication orders and reduced risk of a subsequent assault incident report. Figure 1 shows that a large proportion of total IM medications are ordered within a few hours of presentation, while incident reports peak many hours later. Though IM medications ordered likely overestimate IM medications given, the modest downward slope of the incident curve with respect to length of stay (Figure 1 bottom) suggests that IM medications given still likely outnumber assaults during this period. The overall summative results seen in Table 2 capture a similar story, that IM orders are not associated with a measurable decrease in incidents. Our clinical interpretation is that we did not find evidence that increasing the preemptive use of IM medications would lead to further reduction in assault incident reports.

Regarding the classes of medications used for the management of assault risk, our results suggest that, even under intent-to-treat analysis, antipsychotic medications are associated with a decreased risk of subsequent assault. Viewed in context with the limited finding for IMs, this finding particularly supports the role of scheduled oral antipsychotic medications relative to more invasive IMs in incident prevention. Antipsychotic medications are not benign; side effects can include orthostasis, dystonia, Parkinsonism, metabolic derangements, seizure, and neuroleptic malignant syndrome. The decision of whether to order an antipsychotic is complicated in the acute setting when there is diagnostic uncertainty. That said, if a patient at high risk of assault has a diagnosis where antipsychotics are indicated, these results suggest that antipsychotics may also be helpful in preventing assault.

The findings are limited for benzodiazepines and mood stabilizers. Neither benzodiazepine nor mood stabilizer orders were associated with a subsequent reduction in incident risk in our data, however, this may have more to do with the way in which they are used in our sample. Within our sample, benzodiazepines were seldom used alone for the management of agitation, but more commonly administered in combination with an antipsychotic. When co-administered with an antipsychotic, benzodiazepines did not appear to be associated in any additional reductions in incident risk. These findings appear compatible with consensus guidelines. Hankin et al. (2011) has stated that the goal of agitation management is “to rapidly calm the agitated patient without overly sedating him or her,” and subsequently has suggested that benzodiazepines should not be used alone because they sedate without treating the underlying condition (28).

Mood stabilizers were similarly not found to be associated with reduced incident risk, although their relatively infrequent use limited the analysis. In contrast to antipsychotics, which are administered in 97.7% of visits with incidents, mood stabilizers are prescribed in only 12.6% of such visits. As a result, this analysis may be underpowered to describe a reduction in observed incidents for mood stabilizers. Another explanation may be that mood stabilizers require a longer time to work; clinical trials for valproate and lithium, two common mood stabilizers, have illustrated a 20% reduction in mania scores by 5 days (17,29). By comparison, the median length of stay in the psychiatric emergency room for visits where an incident is reported is 71.2 hours. Median lengths of stay overall are shorter still, 40.4 hours.

As with most observational studies, the primary limitation of this study is in the construction of comparable counterfactual groups (representing what might have happened if circumstances or interventions had been different). The single largest omitted variable is the level of subjective risk as assessed by psychiatric providers, which is not well captured by the durable characteristics of the patient such as age, sex, and violence history only. We attempt to address these concerns by constructing our study longitudinally, and using random effects to correct for inherent differences between patients, though this method is not perfect. Some limitations are inherent to the use of incident report data. It is known that some incidents go unreported. Additionally, using incidents reports as the outcome reduces complex interpersonal events into a binary category (present/absent), which does not account for qualitative differences between the incidents and the parties involved. Important differences exist between the patient with paranoia acting in self-defense, the patient with intellectual disability trying forcefully to leave, and the patient with antisocial traits specifically targeting someone. Future studies would benefit from analyzing specific subsets of incidents.

Safety is a top priority in emergency psychiatry settings. Violence prevention and creation of an environment with “zero harm” are worthwhile goals to pursue. However, the challenge to reduce the prevalence of already rare assaults is complicated. Thoughtful weighing of the risk of in-hospital assault versus the risk of antipsychotic medication exposure is therefore warranted.

## Supporting information

IRB Approval

## Data Availability

Data used in this study are available for research purposes upon request.

## Appendix A.

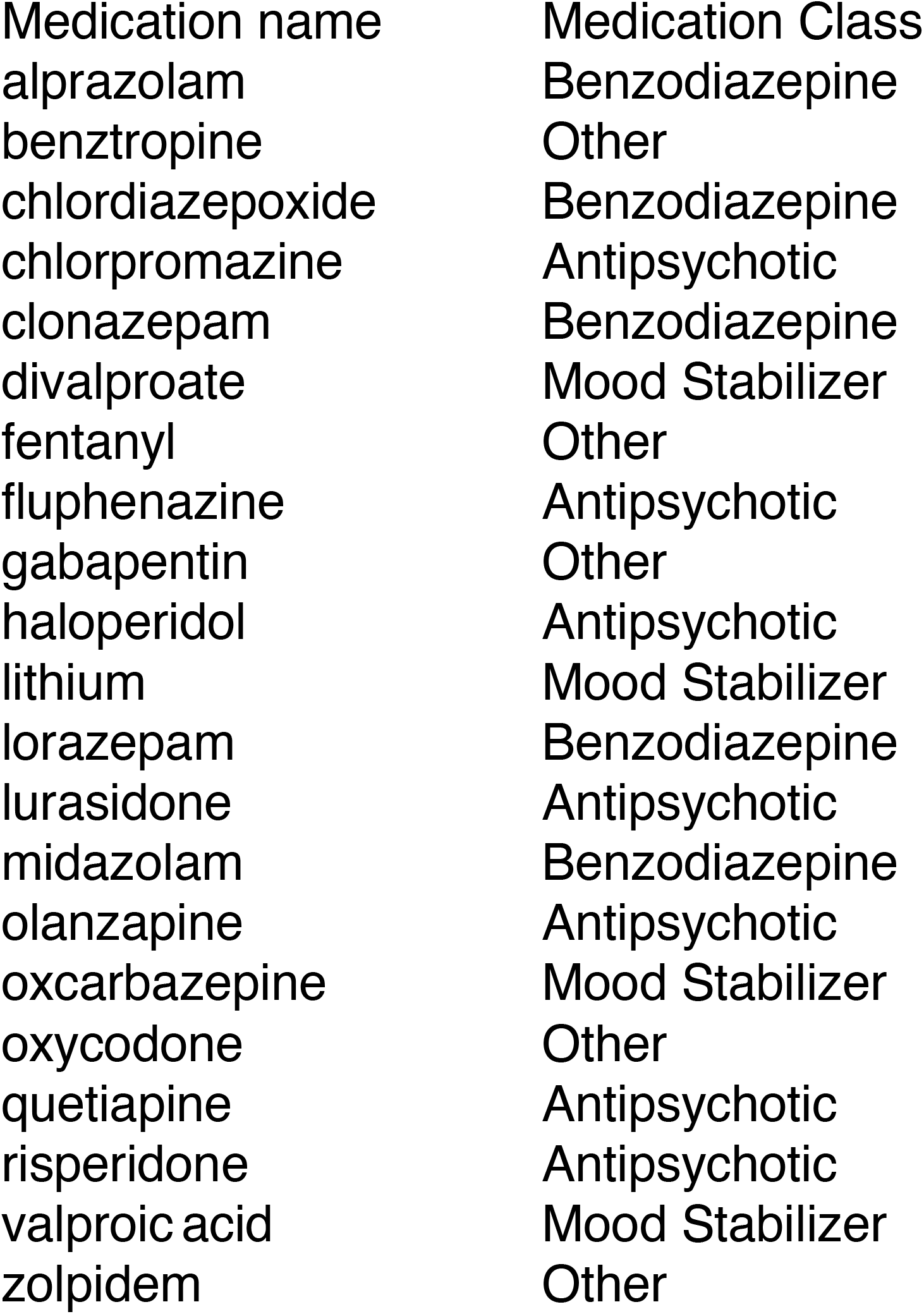

