## Supplementary material for "Medication use and physical assaults in the psychiatric emergency room": IRB Approval

### Columbia University Human Subjects Protocol Data Sheet

#### General Information

| | | | |
|---|---|---|---|
| **Protocol:** | AAAR7763(M00Y03) | **Protocol Status:** | Approved |
| **Effective Date:** | 01/27/2021 | **Expiration Date:** | 01/26/2022 |
| **Originating Department Code:** | | HIC Clinical Informatics (7571419) | |
| **Principal Investigator:** | | Lawrence, Ryan (rel2137) | |
| **From what Columbia campus does this research originate:** | | Medical Center | |
| **Title:** | Studying Patterns of Utilization of Injection Antipsychotic and Assaults In the Psychiatric Emergency Service | | |
| **Protocol Version #:** | | **Abbreviated Title:** | CPEP antipsychotics project |
| **Was this protocol previously assigned a number by an IRB:** | | | No |

**Is the purpose of this submission to obtain a "Not Human Subjects Research" determination?**

No

#### IRB Expedited Determination

**5. Research involving materials (data, documents, records, or specimens) that have been collected, or will be collected solely for nonresearch purposes (such as medical treatment or diagnoses).**

#### Renewal Information

**Enrollment status:**

Closed to further enrollment: study-related procedures ongoing

**Provide any additional information necessary to explain the study status:**

We are not collecting any new data. We have analyzed the data and drafted a manuscript, but have not yet submitted the manuscript for peer review.

**Since the last renewal:**

**Have there been any changes in the relevant literature that would affect the study design or procedures?**

No

**Have there been any interim findings associated with this study?**

No

**Have there been any publications resulting from this study?**

No

**Have any participants been enrolled using the Short Form process?**

No

**Is there a Data Monitoring Committee (DMC), Data Safety Monitoring Board (DSMB), or other monitoring entity for this study?**

No

**Is an annual Progress Report required by the funding organization or coordinating center for this study?**

No

**Does this submission include a modification?**

No

**Has the consent form been revised in this submission?**



Columbia University IRB
( Y3M0 )
Approved for use until: 01/26/2022

No

**Does this submission include a report of a protocol violation?**

No

#### Attributes

**Special review type: Check all that apply or check "None of the Above" box.**

[ ]Review for 45 CFR 46.118 Determination (involvement of human subjects is anticipated but is not yet defined)

[ ]Funding review for Administrative IRB approval (such as for Center or Training Grants)

[x]None of the above

---

**IRB of record information: Will a Columbia IRB be the IRB that is responsible for providing review, approval, and oversight for this study?**

Yes

**Select the most appropriate response:**

**Columbia will be the IRB of record for the study procedures conducted by Columbia researchers (Note: this response will apply to most submissions).**

---

**Is this research part of a multicenter study?**

No

---

**Please indicate if any of the following University resources are utilized:**

[ ] Cancer Center Clinical Protocol Data Management Compliance Core (CPDM)

[ ] CTSA-Irving Institute Clinical Research Resource (CRR)

[ ] CTSA- Irving Institute Columbia Community Partnership for Health (CCPH)

[x] None of the above

#### Background

**Abbreviated Submission:**

**The IRB has an abbreviated submission process for multicenter studies supported by industry or NIH cooperative groups (e.g., ACTG, HVTN, NCI oncology group studies, etc.), and other studies that have a complete stand-alone protocol. The process requires completion of all Rascal fields that provide information regarding local implementation of the study. However, entering study information into all of the relevant Rascal fields is not required, as the Columbia IRBs will rely on the attached stand-alone (e.g., sponsor's) protocol for review of the overall objectives.**

**If you select the Abbreviated Submission checkbox and a section is not covered by the attached stand-alone protocol, you will need to go back and provide this information in your submission.**

---

**Study Purpose and Rationale:**

**Provide pertinent background description with references that are related to the need to conduct this study. If this is a clinical trial, the background should include both preclinical and clinical data. Be brief and to the point.**

[ ] Abbreviated Submission - This information is included in an attached stand-alone protocol. Proceed to the next question

Part of the care program for psychiatry patients at NewYork-Presbyterian Hospital (NYPH)



Columbia University IRB
( Y3M0 )
Approved for use until: 01/26/2022

involves special Emergency Department (ED) units, termed Comprehensive Psychiatric Emergency Program (CPEP) units, designed for psychiatric patients. Unlike the ordinary ED, CPEP allows several-day stays, and is designed and run specifically for psychiatry patients. A significant adverse outcome in the CPEP occurs when patients become ‘agitated’. When patients become highly agitated, the extreme measure must be taken of administering an intramuscular injection of an antipsychotic medication. These medications carry significant side effects and should be considered a last resort. Minimizing unnecessary injections is an important outcome for optimal care. Injection of antipsychotics may also be associated with a number of other adverse outcomes, such as the use of seclusion and restraints and increased number of days of admission.In this study, we aim to uncover the patterns of antipsychotic use in the CUMC Comprehensive Psychiatric Emergency Service (CPEP) in the past few years. We will investigate the trends and demographic breakdown of how antipsychotics are administered, and the association of administration with factors such as previous injections of antipsychotics and information in clinical notes and labs that might indicate agitation. With this information, we will explore whether a predictive model of agitation can be developed, and if this outcome can be avoided through early warning and use of gentler interventions.We also aim to examine cases in which CPEP patients assaulted patients or staff. This data is another lens into the agitation phenotype, and is the ultimate adverse event that is to be avoided. As part of this, we will examine psychiatric drugs (benzodiezapines, oral antipsychotics, and mood stabilizers) and how they affect both incidence of these events and incidence of intramuscular injections of antipsychotics.

---

**Study Design:**

**Describe the methodology that will be used in this study, covering such factors as retrospective vs. prospective data collection, interventional vs. non-interventional, randomized vs. non-randomized, observational, experimental, ethnography, etc.**

[ ] Abbreviated Submission - This information is included in an attached stand-alone protocol. Proceed to the next question

This retrospective chart review will evaluate the medical charts of adult patients in the Columbia CPEP, from January 1, 2010 to December 31, 2019. Patients will be identified based on admit to the CPEP unit any time within the time frame. The following data will be collected: patient demographics; visit length and frequency; severity of illness upon each visit; injection status, clinical notes, and labs from visits within the time frame. Additionally, we propose to incorporate the following variables: medical record number, date/time of presentation to triage, date/time of event, brief description of the event (from Keepsafe Incident report), diagnostic information (ICD-10 codes).

---

**Statistical Procedures:**

**Provide sufficient details so that the adequacy of the statistical procedures can be evaluated including power calculations to justify the number of participants to be enrolled into the study. Definitions of subject terms such as enrolled and accrued as used for Rascal submissions can be found in the Subjects section.**

[ ] Abbreviated Submission - This information is included in an attached stand-alone protocol. Proceed to the next question

Descriptive statistics will be used to summarize information regarding the patient population and overall composition of patient pools receiving and not receiving injections in CPEP. We will compare data using chi-squared test, t-test, and logistic regression models or other generalized linear models. We will also build predictive models using logistic regression or other forms of machine learning. All analysis will be done using the R statistical software, or Python, or STATA. This study is conducted retrospectively, and all available sample is included, therefore no power


Columbia University IRB
( Y3M0 )
Approved for use until: 01/26/2022

analysis is conducted prior to enrollment.

#### Exempt and Expedited

**Is the purpose of this submission to obtain an exemption determination, in accordance with 45CFR46.101(b):**
No

---

**Is the purpose of this submission to seek expedited review , as per the federal categories referenced in 45CFR46.110?**
Yes

**Is the risk of harm to which subjects will be exposed as a result of this research no more than minimal?**
Yes

**Select the category or categories of research into which study procedures fall.**

[ ] Category 1 - Clinical studies of drugs and medical devices only when condition (a) or (b) is met. (a) Research on drugs for which an investigational new drug application (21 CFR Part 312) is not required. (Note: Research on marketed drugs that significantly increases the risks or decreases the acceptability of the risks associated with the use of the product is not eligible for expedited review.) (b) Research on medical devices for which (i) an investigational device exemption application (21 CFR Part 812) is not required; or (ii) the medical device is cleared/approved for marketing and the medical device is being used in accordance with its cleared/approved labeling.

[ ] Category 2 - Collection of blood samples by finger stick, heel stick, ear stick, or venipuncture as follows: (a) from healthy, nonpregnant adults who weigh at least 110 pounds. For these subjects, the amounts drawn may not exceed 550 ml in an 8 week period and collection may not occur more frequently than 2 times per week; or (b) from other adults and children, considering the age, weight, and health of the subjects, the collection procedure, the amount of blood to be collected, and the frequency with which it will be collected. For these subjects, the amount drawn may not exceed the lesser of 50 ml or 3 ml per kg in an 8 week period and collection may not occur more frequently than 2 times per week.

PLEASE NOTE: If blood is collected through an existing catheter, you do not qualify for expedited review under this category.

[ ] Category 3 - Prospective collection of biological specimens for research purposes by noninvasive means. Examples include: (a) hair and nail clippings in a nondisfiguring manner; (b) deciduous teeth at time of exfoliation or if routine patient care indicates a need for extraction; (c) permanent teeth if routine patient care indicates a need for extraction; (d) excreta and external secretions (including sweat); (e) uncannulated saliva collected either in an unstimulated fashion or stimulated by chewing gumbase or wax or by applying a dilute citric solution to the tongue; (f) placenta removed at delivery; (g) amniotic fluid obtained at the time of rupture of the membrane prior to or during labor; (h) supra- and subgingival dental plaque and calculus, provided the collection procedure is not more invasive than routine prophylactic scaling of the teeth and the process is accomplished in accordance with accepted prophylactic techniques; (i) mucosal and skin cells collected by buccal scraping or swab, skin swab, or mouth washings; (j) sputum collected after saline mist nebulization.

[ ] Category 4 - Collection of data through noninvasive procedures (not involving general anesthesia or sedation) routinely employed in clinical practice, excluding procedures involving x-rays or microwaves. Where medical devices are employed, they must be cleared/approved for marketing. (Studies intended to evaluate the safety and effectiveness of the medical device are not generally eligible for expedited review, including studies of cleared medical devices for new indications.) Examples include: (a) physical sensors that are applied either to the surface of the body or at a distance and do not involve input of significant amounts of energy into the


Columbia University IRB
( Y3M0 )
Approved for use until: 01/26/2022

subject or an invasion of the subject=s privacy; (b) weighing or testing sensory acuity; (c) magnetic resonance imaging; (d) electrocardiography, electroencephalography, thermography, detection of naturally occurring radioactivity, electroretinography, ultrasound, diagnostic infrared imaging, doppler blood flow, and echocardiography; (e) moderate exercise, muscular strength testing, body composition assessment, and flexibility testing where appropriate given the age, weight, and health of the individual.

[x]  Category 5 - Research involving materials (data, documents, records, or specimens) that have been collected, or will be collected solely for nonresearch purposes (such as medical treatment or diagnosis). PLEASE NOTE: If extra tissue is being taken during a routine clinical procedure (i.e. additional tissue that is not being taken for diagnostic purposes), you do not qualify for expedited review under this category.

[ ]  Category 6 - Collection of data from voice, video, digital, or image recordings made for research purposes.

[ ]  Category 7 - Research on individual or group characteristics or behavior (including, but not limited to, research on perception, cognition, motivation, identity, language, communication, cultural beliefs or practices, and social behavior) or research employing survey, interview, oral history, focus group, program evaluation, human factors evaluation, or quality assurance methodologies. (NOTE: Some research in this category may be exempt from the HHS regulations for the protection of human subjects. 45 CFR 46.101(b)(2) and (b)(3). This listing refers only to research that is not exempt.)

**Do all procedures fall into one or more of the categories listed above?**

Y

**NOTE:  This project appears to be eligible for expedited review.**

#### Funding

**Is there any external funding or support that is applied for or awarded, or are you the recipient of a gift, for this project?**

No

#### Locations

| Location Type | Facility Name | Domestic or International | Geographic Location | Local IRB Ethics Approval | Local Site Approval |
|---|---|---|---|---|---|
| NewYork-Presbyterian Hospital @ Columbia | 601 West 168th Street | | | | |

#### Personnel

| UNI/Phone | Name | Role | Department | Edit/View | Obtaining Informed Consent |
|---|---|---|---|---|---|
| rel2137<br><br>603-997-6797 | Lawrence, Ryan | Principal Investigator | PSY Clinical Programs (754520X) | Edit | N |



Columbia University IRB
( Y3M0 )
Approved for use until: 01/26/2022

| UNI/Phone | Name | Role | Department | Edit/View | Obtaining Informed Consent |
|---|---|---|---|---|---|
| | **Roles and Experience:** Ryan Lawrence is the Director of the Comprehensive Psychiatric Emergency Program, the clinical area where all subjects received treatment. With David Vawdrey transitioning to a smaller role on this protocol, Ryan will take over as Principal Investigator. Ryan has experience analyzing clinical datasets and publishing results. As PI he will assume responsibility for storing the dataset on an encrypted device, and oversee the analysis of the data (with input from co-investigators) and the writeup. | | | | |
| dkv2101<br>212-305-9801 | Vawdrey, David | Investigator | DBM DBMI (756700X) | Edit | N |
| | **Roles and Experience:** David Vawdrey was the original PI on the project and has expertise in analyzing hospital data. He has moved on to other projects, so has requested to transition to an Investigator role. | | | | |
| mo2707<br>347-514-3670 | Oberhardt, Matthew | Investigator | OBG Research (752830X) | Edit | N |
| xsl2101<br>646-774-6138 | Luo, Sean | Investigator | PSY Substance Use Disorders (754430X) | Edit | Y |
| | **Roles and Experience:** Dr. Luo is the originator of the project and a doctor experienced with the CPEP operation, who will provide scientific, methodological, and clinical expertise. He is Assistant Professor of Clinical Psychiatry, Department of Psychiatry, Columbia University. | | | | |
| yg2687<br>646-774-6300 | Gao, Yihe | Investigator | PSY Central Department (754320X) | View | N |
| | **Roles and Experience:** Dr Gao has a MD/PhD in economics and is currently a psychiatry resident at Columbia. She is joining the project to help with data analysis. | | | | |

**Training and COI**

**The PI must ensure that each individual that is added as personnel has met the training requirements for this study (http://www.cumc.columbia.edu/dept/irb/education/index.html). For help identifying which research compliance trainings you may be required to take, visit the Research Compliance Training Finder.**

| UNI | Name | COI | HIPAA | HSP (CITI) | Research with Minors (CITI) | FDA-Regulated Research (CITI) | S-I | CRC | Good Clinical Practice (GCP) | GCP - Third-party tracking | GCP Refresher | Genetic Research Consent |
|---|---|---|---|---|---|---|---|---|---|---|---|---|
| rel2137 | Lawrence, Ryan | 12/01/2020 | 06/11/2016 | 03/20/2019 | | 03/20/2019 | 06/11/2016 | | | | | |
| dkv2101 | Vawdrey, David | 02/04/2020 | 08/08/2007 | 02/03/2020 | 01/09/2016 | 01/09/2016 | | | | | | |
| mo2707 | Oberhardt, Matthew | 08/27/2020 | 09/16/2016 | 01/13/2020 | 09/02/2016 | 09/02/2016 | | | 01/13/2020 | | | |
| xsl2101 | Luo, Sean | 10/14/2020 | 07/09/2014 | | | | | | | | | |
| yg2687 | Gao, Yihe | 03/09/2020 | 03/05/2020 | 03/05/2020 | | | | | | | | |

#### Departmental Approvers

Electronic Signature: Ryan Lawrence (754520X) - Principal Investigator Date: 01/22/2021



Columbia University IRB
( Y3M0 )
Approved for use until: 01/26/2022

#### Privacy & Data Security

**Indicate the methods by which data/research records will be maintained or stored (select all that apply):**

[ ]Hardcopy (i.e., paper)

[x]Electronic

**Where will the data be stored?**

Y

[ ]On a System

[x]On an Endpoint

**Identify what type of endpoint will be used (select all that apply):**

[x]Desktop Computer

[x]Laptop Computer

[ ]Mobile Device

[ ]Other

---

**Does this study involve the receipt or collection of Sensitive Data?**

Yes

**If any Sensitive Data is lost or stolen as part of your research protocol, you must inform both the IRB and the appropriate IT Security Office (CUMC IT Security if at CUMC; CUIT if at any other University campus).**

**What type of Sensitive Data will be obtained or collected? Select all that apply:**

[x]Personally Identifiable Information (PII), including Social Security Numbers (SSN)

**Will Social Security Numbers (SSNs) be collected for any purpose?**

No

[x]Protected Health Information (PHI), including a Limited Data Set (LDS)

**If any PHI is lost or stolen, you must inform both the IRB and the Office of HIPAA Compliance.**

**Indicate plans for secure storage of electronic sensitive data: check all that apply**

[ ]Sensitive data will not be stored in electronic format

[ ]Sensitive data will be stored on a multi-user system

[x]Sensitive data will be stored on an encrypted endpoint

**By Selecting an Endpoint Device and approving this protocol for submission to the IRB, the PI is attesting that the device and any removable media that may be used have been or will be registered and/or will be maintained in compliance with the University's Information Security Charter and all related policies. It is important that this information is updated, during the course of the study, as new devices are added.**

---

**Provide a description of how the confidentiality of study data will be ensured, addressing concerns or protections that specifically relate to the data storage elements identified above (e.g. hard copy, electronic, system, and/or endpoint):**

The data that are extracted from the CDW containing patient and user identifiers will be kept on secure, password protected, encrypted, single user endpoints, including encrypted laptops of the investigators. Authorization to access research data is strictly limited to the project personnel, and they will have signed the medical center's confidentiality



Columbia University IRB
( Y3M0 )
Approved for use until: 01/26/2022

agreement. Protected health information will be discarded once analysis is complete.

**If your project is not NIH funded, has a Certificate of Confidentiality (CoC) been requested for this research?**
No

---

**Provide a description of the protections in place to safeguard participants' privacy while information is being collected:**
Patient records will only be accessed by study personnel and only study specific parameters will be reviewed.

#### Procedures

**Is this project a clinical trial?**
No
**Is this project associated with, or an extension of, an existing Rascal protocol?**
No
**Do study procedures involve any of the following?**

**Analysis of existing data and/or prospective record review**
Yes
**Audio and/or video recording of research subjects**
No
**Behavioral Intervention?**
No
**Biological specimens (collection or use of)**
No
**Cancer-related research**
No
**Drugs or Biologics**
No
**Future use of data and/or specimens**
No
**Genetic research**
No
**Human embryos or human embryonic stem cells**
No
**Imaging procedures or radiation**
No
**Medical Devices**
No
**Surgical procedures that would not otherwise be conducted or are beyond standard of care**
No

**Will any of the following qualitative research methods be used?**

**Survey/interview/questionnaire**
No
**Systematic observation of public or group behavior**
No
**Program evaluation**


Columbia University IRB
( Y3M0 )
Approved for use until: 01/26/2022

No

**Will any of the following tests or evaluations be used?**

**Cognitive testing**

No

**Educational testing**

No

**Non-invasive physical measurements**

No

**Taste testing**

No

**Is there an external protocol that describes ALL procedures in this study?**

No

**Please describe ALL study procedures in detail.**

**NOTE: Be sure to detail all of the procedures above to which a "yes" response was selected. Also detail any additional procedures that may or may not fall into the categories listed above.**

This is a retrospective chart review evaluation.

#### Analysis of Existing Data and/or Prospective Record Review

**Indicate whether the data that will be collected or utilized for the proposed study are in existence as of the current IRB submission date.**

All of the data are in existence

**Provide the date range of the existing data, documents, or records (e.g., medical charts, school records, census data)**

Beginning Date: 01/01/2010
End Date: 12/31/2019

**Note that end dates beyond the initial IRB Protocol submission date or future requests for a date parameter extension beyond the provided end date may require informed consent and HIPAA Authorization to be obtained from subjects.**

---

**Data will be obtained from (select all that apply):**

[x] Columbia and/or NYP (e.g., departmental databases/systems, patient charts, Eclipsys, WebCIS, administrative/billing records, etc.)

**Select all that apply:**

[x] Data to be analyzed were or will be collected for clinical care


Columbia University IRB
( Y3M0 )
Approved for use until: 01/26/2022

[ ] Data to be analyzed were or will be collected for nonresearch purposes other than for clinical care (e.g., student records, class evaluation, administrative records, etc.,)

[ ] Data originate from an IRB approved protocol

[ ] Other

[ ] Outside Columbia and/or NYP:

---

**Will a member of the research team be abstracting data directly from source documents?**

Yes

**If there is a data abstraction document/spreadsheet, attach it to the submission to complete study records. Though the IRB does not approve these documents, for reference purposes they are extremely helpful in understanding the scope of the proposed data collection.**

**Select the applicable responses:**

[x] The data, documents, or records to be reviewed/abstracted are those to which a member of the research team has legitimate access for non-research purposes (e.g., departmental patient database, physicians' patient clinical records, student records).

[ ] Special authorization is necessary to review the records as the research team does not have access to the data, and a request will be or has been made to access the data.

---

**If any existing data was obtained from a prior research study, was any member of the current research team involved (e.g., obtained consent, performed study procedures, conducted data analysis) in the project or procedures that collected and/or used identifiable information?**

N/A

---

**Indicate the manner in which the existing data and/or the records to be reviewed prospectively will be collected or received:**
**(Select all that apply. At least one must be selected.)**

[x] Contains direct identifiers (e.g., name, MRN, date of birth)
[ ] Coded and the research team has the key and can link the data to direct identifiers
[ ] Coded and the research team does not have access to the key to link data to direct identifiers
[ ] Prior to the receipt of the data by the research team submitting this protocol, the identifiers will be removed and


Columbia University IRB
( Y3M0 )
Approved for use until: 01/26/2022

no link will remain.

[ ] The information was originally or will be collected without identifiers

**If data are collected or received at any point in time with direct identifiers or linked to identifiers, then the data are considered to be identifiable, and the requirements for Informed Consent (or a waiver, if applicable) and HIPAA Authorization (or a waiver, if applicable) apply. The necessary information will need to be included in the respective sections of the submission.**

#### Recruitment And Consent

##### Recruitment:

**Will you obtain information or biospecimens for purposes of screening or determining eligibility?**

No

**Describe how participants will be recruited:**

This is a retrospective chart review.

**Select all methods by which participants will be recruited:**

[x] Study does not involve recruitment procedures
[ ] Person to Person
[ ] Radio
[ ] Newspapers
[ ] Direct Mail
[ ] Website
[ ] Email
[ ] Television
[ ] Telephone
[ ] Flyer/Handout
[ ] Newsletter/Magazine/Journal
[ ] ResearchMatch
[ ] CUMC RecruitMe

**Additional Study Information: Please add a description of your study as you would like it to be displayed on the RecruitMe website.**

---

##### Informed Consent Process:

**Informed Consent Process, Waiver or Exemption: Select all that apply**

[ ] Informed consent with written documentation will be obtained from the research participant or appropriate representative.

[ ] Informed consent will be obtained but a waiver of written documentation of consent (i.e., agreement to participate in the research without a signature on a consent document) is requested.


Columbia University IRB
( Y3M0 )
Approved for use until: 01/26/2022

[x]  A waiver of some or all elements of informed consent (45 CFR 46.116) is requested.

**Waiver of consent is applicable to:**

The study in its entirety

**Select the applicable situation:**

[x]This study qualifies for a waiver or alteration of consent as the following criteria are met in this study (provide justification for EACH of these criteria):

**(1)The research involves no more than minimal risk to the subjects**

**Provide justification:**

We will analyze the data that is already collected as part of clinical care, and will not report any individual outcomes. All results will be in aggregate. The only foreseeable risk is the risk of breech of confidentiality but we have planned appropriate safeguards to preserve the privacy of participants.

**(2) The waiver or alteration will not adversely affect the rights and welfare of the subjects**

**Provide justification:**

Data is not reported as individual outcomes. The reports will not affect the individuals whose data will be analyzed as they will not be identifiable.

**(3) The research could not practicably be carried out without the waiver or alteration**

**Provide justification:**

Originally we designed this as a retrospective research study, therefore it was impractical at the outset to obtain consent from all of the patients, especially because we are evaluating historical data and many patients are no longer treated at the organization. Following some preliminary analyses of our data earlier this year, we realized there are some additional variables that would be essential to our analysis, and we realized that our dataset was becoming outdated (data were collected only through 2017). In this modification we are requesting a one time extension in order to make our dataset more current (using data through 2019).

**(4) Whenever appropriate, the subjects will be provided with additional pertinent information after participation**

**Provide justification:**

Not applicable to this research.

**(5)  If the research involves using identifiable private information or identifiable biospecimens, the research could not practicably be carried out without using such information or biospecimens in an identifiable format**

****This waiver criterion does not apply if your research was initially approved before January 21, 2019, which is the general compliance date for the revised regulations at 45CFR46 subpart A.**

**Provide justification:**

This research requires using identifiable private information because we are matching Keepsafe Reports with information in the electronic health record. Without identifiable private information



Columbia University IRB
( Y3M0 )
Approved for use until: 01/26/2022

(especially name, mrn) the matching would not be possible.

[ ]This study qualifies for waiver or alteration of consent involving public benefit and service programs as the following criteria are met for this study (provide justification for EACH of these criteria):

[ ] Planned Emergency Research with an exception from informed consent as per 21 CFR 50.24.

[ ] This is exempt research.

---

**Subject Language**

Language of subjects is unknown/irrelevant (e.g., record reviews, mass mailing of surveys)

---

**Capacity to Provide Consent:**

**Do you anticipate using surrogate consent or is research being done in a population where capacity to consent may be questionable?**

No

#### Research Aims & Abstracts

**Research Question(s)/Hypothesis(es):**

The following research questions will be addressed: 1) What are the characteristics of patients who end up getting antipsychotic intramuscular injections in CPEP?

2) What are the early clinical markers of 'agitation'? 3) What associations do antipsychotic injections have with increased length of stay and other adverse outcomes? 4) What *causal* associations do antipsychotic injections have with increased length of stay and other adverse outcomes (if possible to assess)? 5) Can we predict intramuscular injections using pre-injection data? 6) What are the early clinical markers and characteristics of CPEP patients who assault other patients or staff, and how does this relate to antipsychotic injections? 7) How do psychiatric drug regimens affect likelihood of patients assaulting other patients or staff?

---

**Scientific Abstract:**

Part of the care program for psychiatry patients at NewYork-Presbyterian Hospital (NYPH) involves special Emergency Department (ED) units, termed Comprehensive Psychiatric Emergency Program (CPEP) units, designed for psychiatric patients. Unlike the ordinary ED, CPEP allows several-day stays, and is designed and run specifically for psychiatry patients. A significant adverse outcome in the CPEP occurs when patients become ‘agitated’. When patients become highly agitated, the extreme measure must be taken of administering an intramuscular injection of an antipsychotic medication. These medications carry significant side effects and should be considered a last resort. Minimizing unnecessary injections is an important outcome for optimal care. Injection of antipsychotics may also be associated with a number of other adverse outcomes, such as the use of seclusion and restraints and increased number of days of admission.In this study, we aim to uncover the patterns of antipsychotic use in the CUMC Comprehensive Psychiatric Emergency Service (CPEP) in the past few years. We will investigate the trends and demographic breakdown of how antipsychotics are administered, and the association of administration with factors such as previous injections of antipsychotics and information in clinical



Columbia University IRB
( Y3M0 )
Approved for use until: 01/26/2022

notes and labs that might indicate agitation. With this information, we will explore whether a predictive model of agitation can be developed, and if this outcome can be avoided through early warning and use of gentler interventions. .We will also examine the likelihood of a CPEP patient assaulting other patients or staff as a final adverse outcome and derivative of the agitation phenotype. We will explore if our predictive model can also address assaults, and the roles that psychiatric drugs (e.g., Benzodiezapines and mood stabilizers) play in modulating agitation and assaults.

---

**Lay Abstract:**

Part of the care program for psychiatry patients at NewYork-Presbyterian Hospital (NYPH) involves special Emergency Department (ED) units, termed Comprehensive Psychiatric Emergency Program (CPEP) units, designed for psychiatric patients. Unlike the ordinary ED, CPEP allows several-day stays, and is designed and run specifically for psychiatry patients. A significant adverse outcome in the CPEP occurs when patients become 'agitated'. When patients become highly agitated, the extreme measure must be taken of administering an intramuscular injection of an antipsychotic medication. These medications carry significant side effects and should be considered a last resort. Minimizing unnecessary injections is an important outcome for optimal care. Injection of antipsychotics may also be associated with a number of other adverse outcomes, such as the use of seclusion and restraints and increased number of days of admission.In this study, we aim to uncover the patterns of antipsychotic use in the CUMC Comprehensive Psychiatric Emergency Service (CPEP) in the past few years. We will investigate the trends and demographic breakdown of how antipsychotics are administered, and the association of administration with factors such as previous injections of antipsychotics and information in clinical notes and labs that might indicate agitation. With this information, we will explore whether a predictive model of agitation can be developed, and if this outcome can be avoided through early warning and use of gentler interventions. We will also examine the likelihood of a CPEP patient assaulting other patients or staff as a final adverse outcome and derivative of the agitation phenotype. We will explore if our predictive model can also address assaults, and the roles that psychiatric drugs (e.g., Benzodiezapines and mood stabilizers) play in modulating agitation and assaults.

#### Risks, Benefits & Monitoring

**Abbreviated Submission:**

**The IRB has an abbreviated submission process for multicenter studies supported by industry or NIH cooperative groups (e.g., ACTG, HVTN, NCI oncology group studies, etc.), and other studies that have a complete stand-alone protocol. The process requires completion of all Rascal fields that provide information regarding local implementation of the study. However, entering study information into all of the relevant Rascal fields is not required, as the Columbia IRBs will rely on the attached stand-alone (e.g., sponsor's) protocol for review of the overall objectives. .**

**If you select the Abbreviated Submission checkbox and a section is not covered by the attached stand-alone protocol, you will need to go back and provide this information in your submission.**

---

**Potential Risks:**



Columbia University IRB
( Y3M0 )
Approved for use until: 01/26/2022

**Provide information regarding all risks to participants that are directly related to participation in this protocol, including any potential for a breach of confidentiality. Risks associated with any of the items described in the Procedures section of this submission should be outlined here if they are not captured in a stand-alone protocol. Risks of procedures that individuals would be exposed to regardless of whether they choose to participate in this research need not be detailed in this section, unless evaluation of those risks is the focus of this research. When applicable, the likelihood of certain risks should be explained and data on risks that have been encountered in past studies should be provided.**

[ ] Abbreviated Submission - This information is included in an attached stand-alone protocol. Proceed to the next question

The data used in this study will contain patient and provider identifiers. Provider identifiers will include their name and their unique identifier (the username they use to access the electronic health records). Patient identifiers will include the patient's medical record number only. Because this data includes protected health information (PHI), and addresess (PII), there is a potential risk of breech of confidentiality. However, we have planned appropriate safeguards to avoid this risk. All data will be only stored on encrypted endpoint devices that are only accessible to the research team (which are listed as personnel for this protocol). We will use the same encryption technology that has been approved by CUIT and is currently in use by our CUIT approved devices. All results will be reported in aggregate and with no identifiers.

---

**Potential Benefits:**

**Provide information regarding any anticipated benefits of participating in this research. There should be a rational description of why such benefits are expected based on current knowledge. If there is unlikely to be direct benefit to participants/subjects, describe benefits to society. Please note that elements of participation such as compensation, access to medical care, receiving study results, etc. are not considered benefits of research participation.**

[ ] Abbreviated Submission - This information is included in an attached stand-alone protocol. Proceed to the next question

Outcomes of the current study can be beneficial in several ways.

First, they can provide us with insight as to what factors contribute to and are predictive of agitation and violence of psychiatric patients. The association between intramuscular injection of antipsychotic medications and various adverse outcomes (e.g., increased length of stay and assaults) will be explored, which should help contextualize the injections in the broader scope of hospital care, and help determine if reducing injections alone is a worthwhile goal.

Next, the resulting description of CPEP dynamics at Columbia, alongside the predictive model of who is likely to get injected, will lay a groundwork for future interventions through which intramuscular injections may be avoided. This will result in less expensive and higher quality care, thereby increasing the overall value of the care provided at our institution.

Finally, the insights acquired through this study can be disseminated in presentations and publications, which would potentially help other institutions in planning and implementation of similar interventions at their institutions.

---

**Alternatives:**

**If this research involves an intervention that presents greater than minimal risk to participants, describe available alternative interventions and provide data to support their efficacy and/or availability. Note, participants always have the option not to participate in research.**

[ ] Abbreviated Submission - This information is included in an attached stand-alone protocol. Proceed to the next question

This study is conducted retrospectively therefore participants will not have an alternative to participation.

---

**Data and Safety Monitoring:**

**Describe how data and safety will be monitored locally and, if this is a multi-center study, how data and safety**


Columbia University IRB
( Y3M0 )
Approved for use until: 01/26/2022

**will be monitored across sites as well.**

[ ]  Abbreviated Submission - This information is included in an attached stand-alone protocol. Proceed to the next question

We will store the data only on encrypted endpoint devices. Wherever applicable, we will replace the personal identifiers with research identifiers that are randomly generated. A mapping between the protected identifiers and research identifiers will be kept in a file separate from that being analyzed, and will also be stored only on encrypted endpoint devices. We will use the same encryption technology that is approved by CUIT and we currently use on our CUIT approved devices.

#### Subjects

**Unless otherwise noted, the information entered in this section should reflect the number of subjects enrolled or accrued under the purview of Columbia researchers, whether at Columbia or elsewhere.**

**Target enrollment:**

25,000

**Number enrolled to date:**

13,816

**Number enrolled since the last renewal or, if this is the first renewal, since the initial approval:**

2,624

**Number anticipated to be enrolled in the next approval period:**

0

**Does this study involve screening/assessment procedures to determine subject eligibility?**

No

**Of the number of subjects enrolled, or the number accrued for interventional studies with a screening process:**

**How many remain on the study?**

0

**How many are off study?**

0

**How many completed the study?**

0

**Have any withdrawn of their own initiative?**

No

**Have any been removed by PI?**

No

**Have any been lost to follow-up?**

No

**Have any died while on study?**

No

**Have any subject complaints been received?**

No

**Is this a multi-center study?**

No

**Does this study have one or more components that apply to a subset of the overall study population (e.g. Phase 1/2, sub-studies)?**

No

**Of the number enrolled, or the number accrued for interventional studies with a screening process, indicate:**

**Population Gender**


Columbia University IRB
( Y3M0 )
Approved for use until: 01/26/2022

| Females | Males | Non Specific |
|---|---|---|
| 33% | 66% | 1% |

**Population Age**

| 0-7 | 8-17 | 18-65 | >65 | Non Specific |
|---|---|---|---|---|
| 0% | 0% | 80% | 20% | 0% |

**Population Race**

| American Indian/Alaskan Native | Asian | Native Hawaiian or Other Pacific Islander | Black or African American | White | More than One Race | Non-Specific |
|---|---|---|---|---|---|---|
| 0% | 10% | 0% | 20% | 30% | 40% | 0% |

**Population Ethnicity**

| Hispanic or Latino | Not Hispanic or Latino | Non-Specific |
|---|---|---|
| 60% | 40% | 0% |

**Vulnerable Populations as per 45 CFR 46:**

**Will children/minors be enrolled**

No

**Will pregnant women/fetuses/neonates be targeted for enrollment?**

No

**Will prisoners be targeted for enrollment?**

No

**Other Vulnerable Populations:**

[ ]Individuals lacking capacity to provide consent

[ ]CU/NYPH Employees/Residents/Fellows/Interns/Students

[ ]Economically disadvantaged

[ ]Educationally disadvantaged

[ ]Non-English speaking

[x]Other Vulnerable populations

These are psychiatric patients, so they may be mentally impaired and unable to fully provide consent. Since this is a retrospective chart review there should be no harm to the patients and nothing related to their care will be affected.

[ ]None of the Populations listed above will be targeted for Enrollment

**Subject Population Justification:**

By necessity of the project, we are dealing with psychiatric patients, many of whom will not be able to provide consent due to psychiatric conditions. However, we are dealing with retrospective data and are seeking waived need for consent, and the harms are low.

**Does this study involve compensation or reimbursement to subjects?**

No

#### Attached HIPAA Forms

| Number | Type | Title | Status |
|---|---|---|---|
| AAAO0553 | B | CPEP form B | Approve |



Columbia University IRB
( Y3M0 )
Approved for use until: 01/26/2022

#### Documents

| Archived | Document Identifier | Document Type | File Name | Active | Stamped | Date Attached | Created By |
|---|---|---|---|---|---|---|---|
| No | PI transfer email | Email/Communication/Message | PI transfer.pdf | Y | No | 02/22/2020 | Ryan Lawrence (rel2137) |
| No | Billing Approval | Other | Billing Approval (AAAR7763).pdf | Y | No | 07/23/2020 | Ashley Halinski (ah3675) |
| No | Sean Luo CITI training | Other | Sean Luo CITI.pdf | Y | No | 06/06/2020 | Ryan Lawrence (rel2137) |
| No | Sean Luo CITI | Other | Sean_Luo_form.pdf | Y | No | 09/04/2019 | Matthew Oberhardt (mo2707) |

#### Tasks

| Section | Task | Required for submission | Completed | Created By | Date Created |
|---|---|---|---|---|---|
| Personnel | Previous IRB correspondence has yet to be addressed: Please update the CITI profile for Sean Luo to reflect his affiliation with Columbia University. Note the training attached in RASCAL lists his affiliated institution as NYPSI. Training will not reflect in RASCAL until Sean's CITI affiliation is listed as Columbia University. | Yes | No | Carri-Ann Gay (cg2618) | 2021-01-25 10:55:10.0 |



Columbia University IRB
( Y3M0 )
Approved for use until: 01/26/2022
